# Smartphone Postural Sway and Pronator Drift tests as Measures of Neurological Disability

**DOI:** 10.1101/2024.11.20.24317196

**Authors:** Michael Calcagni, Peter Kosa, Bibi Bielekova

## Abstract

The COVID-19 pandemic and increased demands for neurologists have inspired the creation of remote, digitalized tests of neurological functions. This study investigates two tests from the Neurological Functional Tests Suite (NeuFun-TS) smartphone application, the “Postural Sway” and “Pronator Drift” tests. These tests capture different domains of postural control and motoric dysfunction in healthy volunteers (n=13) and people with neurological disorders (n=68 relapsing-remitting multiple sclerosis [MS]; n=21 secondary progressive MS; n=23 primary progressive MS; n=13 other inflammatory neurological diseases; n=21 non-inflammatory neurological diseases; n=4 clinically isolated syndrome; n=1 radiologically isolated syndrome). Smartphone accelerometer data was transformed into digital biomarkers, which were filtered in the training cohort (∼80% of subjects) for test-retest reproducibility and correlations with subdomains of neurological examinations and validated imaging biomarkers. The independent validation cohort (∼20%) determined whether biomarker models outperformed the best single digital biomarkers. Postural sway acceleration magnitude in the eyes closed and feet together stance demonstrated the highest reliability (ICC=.706), strongest correlations with age (Pearson r<=.82) and clinical and imaging outcomes (r<=.65, p<0.001) and stronger predictive value for sway-relevant neurological disability outcomes than models that aggregated multiple biomarkers (coefficient of determination R^2^=.46 vs .38). The pronator drift test only captured cerebellar dysfunction, had less reproducible biomarkers, but provided additive value when combined with postural sway biomarkers into models predicting global scales of neurological disability. In conclusion, a simple 1-minute postural sway test accurately measures body oscillations that increase with natural aging and differentiates them from abnormally increased body oscillations in people with neurological disabilities.

## Introduction

### In the wake of the Coronavirus pandemic, the need for remote patient care is clear

Communication technologies have opened the doors to telemedicine, but hands-on aspects of clinical evaluation remain to be replicated. In particular, neurological examinations could benefit from a remote alternative. Full neurological exams are difficult to complete in busy clinical settings and can be subjective in nature. Consequently, functional tests such as timed-walk tests or the 9-hole peg test have been performed in clinical research to bolster clinician-driven disability scales. Digitalization of these tests offers advantages and some tests have been aggregated into suites adapted for smartphones and tablets [1–5]. To our knowledge, the Neurological Function Tests Suite (NeuFun-TS), which is comprised of sixteen smartphone tests measuring various neurological domains, provides the most comprehensive assessment of nervous system functionality. NeuFun-TS outcomes correlate with Multiple Sclerosis (MS) disability scales, brain MRI data, and targeted subdomains of gold-standard neurological examinations [6, 7]. This study examines the NeuFun-TS Postural Sway and Pronator Drift tests.

The Postural Sway test evaluates human upright stance, which is fundamentally unstable. The brain must continuously integrate multisensory input from visual, vestibular, and proprioceptive systems to maintain balance. Since the mid-20^th^ century, postural sway tests have been performed to gain insight into these subsystems. In a typical test, a subject stands in different positions for a short time (e.g., 30 seconds); positions increase in difficulty by inhibiting sensory input and/or increasing stance instability (i.e., standing on foam, closing eyes) while a force plate or body-worn accelerometer records sway data.

The Pronator Drift test examines sustained supination of outstretched arms (i.e., palms facing upwards) in the absence of visual stimulus. This test records involuntary movements, such as tremors, and stereotypical pronation, elbow flexion and downward drift, which may reveal subclinical motoric dysfunction.

Both tests use a smartphone-embedded accelerometer. While force plates were the gold-standard for measuring postural sway, accelerometers mimic force plate measurements with comparable reliability [8]. Furthermore, accelerometry-derived measurements can differentiate between control groups and people with Parkinson’s Disease, Huntingdon’s Disease, MS, and other neurological diseases [9–11]. Similarly, accelerometry quantifies hand tremor and sway to distinguish healthy subjects from people with Parkinson’s Disease, acute ischemic stroke, and Essential Tremor [12, 13].

However, while qualitatively distinguishing “healthy volunteers” (HV) from people with identified neurological disease is important, *disability exists on a spectrum*; more useful digital outcomes quantify neurological disability in a subject with longitudinal accuracy to measure neurological functions agnostically and longitudinally, capable of measuring both disability progression and therapeutic effect. Towards this purpose, some studies correlated postural sway measurements with neurological-disability scales, such as the Expanded Disability Status Scale (EDSS) [14]. While EDSS serves as a progression outcome in MS clinical trials, this ordinal scale (ranging from 0-10) is insensitive for most research applications that use smaller patient cohorts because a minority actually progress on EDSS yearly; e.g., in the ORATORIO clinical trial only 39.3% of primary-progressive MS (PPMS) patients randomized to placebo progressed over 4.2 years of trial duration [15], achieving annual progression rates under 10%. Thus, granular scales like CombiWISE (continuous scale from 0-100) and NeurEx^TM^ (continuous scale from 0 to theoretical maximum of 1349) that strongly correlate with EDSS but measure disability progression in 6-12 months are better suited for research applications, including differentiating disability in specific subdomains of the neurological examination [16, 17].

This study analyzes accelerometry data from the NeuFun-TS Postural Sway and Pronator Drift tests to generate digital biomarkers, assess their test-retest reliability and their correlation with gold-standard clinical outcomes and validated semiquantitative imaging biomarkers. We also explored whether aggregating reliable digital biomarkers into machine-learning (ML) optimized models provide greater clinical value than the best single biomarkers from each test.

## Materials and Methods

### Participants

This study was approved by the Institutional Review Board of the National Institutes of Health (NIH). All data were collected under protocols: Targeting Residual Activity by Precision, Biomarker-Guided Combination Therapies of Multiple Sclerosis (clinicaltrials.gov identifier NCT03109288) and Comprehensive Multimodal Analysis of Neuroimmunological Diseases of the Central Nervous System (NCT00794352). All participants signed paper or digital informed consent and provided their sex, age, height, and weight. After unblinding diagnostic categories, the cohort consisted of people with MS (n=112), Other Inflammatory Neurological Diseases (OIND, n=13), Non-Inflammatory Neurological Diseases (NIND, n=21), Clinically Isolated Syndrome (CIS, n=4), Radiologically Isolated Syndrome (RIS, n=1) and 13 healthy volunteers (HV). Apart from 7 self-declared HV that participated in the “smartphone only substudy cohort”, the remaining 157 study subjects received full neurological and physical examinations, laboratory testing (blood and cerebrospinal fluid to make/confirm diagnosis) and brain MRI within 1–48 h of NeuFun-TS testing (Supplementary Tables 1-2).

### Clinical and imaging measurements related to postural sway and pronator drift

Each neurological examination performed by an MS-trained clinician lasted approximately 30-60 minutes and was documented in the NeurEx™ app [16], which automatically computes neurological disability scales and functional subdomains. Based on domain expertise, we selected NeurEx^TM^ panel scores of neurological functions that contribute to postural sway, including “Stance & Gait” (panel 16), “Cerebellar Dysfunction” (Lower extremities subdomain of panel 12), and “Proprioceptive Dysfunction” (Lower extremities subdomain of panel 14). The square root of the sum of these scores computed the “NeurEx^TM^ Postural Sway” subpanel. Similarly, for the Pronator Drift analysis we selected the pertinent subsystem scores separately for each hand; we derived upper extremity “Motoric Dysfunction” (sums panel 8: muscle strength, panel 10: reflexes and panel 7: spasticity), “Cerebellar Dysfunction” (panel 12 for a specific hand), and “Proprioceptive Dysfunction (panel 14 for a specific hand). We summed these subsystem scores for both hands to compute “NeurEx^TM^ Pronator Drift”.

Clinical grade magnetic resonance imaging (MRI) of the brain and upper cervical spinal cord (i.e., axial and sagittal cuts extended to C5 level) was performed following procedures detailed in previous papers [18, 19]. We semi-quantitatively graded the extent of atrophy and number of focal lesions (“lesion load”) separately in the brainstem, cerebellum, and medulla/upper cervical spinal cord using a previously published grading protocol; these measurements have proven to be important in determining physical disability [18, 19].

Data were collected by investigators blinded to other measurements, uploaded to the research database each week following patient visits, quality controlled during weekly meetings, and locked from subsequent data modifications.

### NeuFun-TS tests description

NeuFun-TS tests were developed using Kotlin and Java within Android Studio. NeuFun-TS is distributed as an Android package and test results are uploaded to a private database hosted on Google Firebase. Results are linked to an alphanumeric code to maintain privacy. The test operates on Android (Versions 11, 12) and displays graphics optimized for Google Pixel XL/2XL.

NeuFun-TS test subjects are provided with a Google Pixel smartphone that has an attached hand strap; they also receive an Auro Lounger Universal Adjustable Neck Mount (phone harness) to be used for the Postural Sway test. For each test in the app suite, when the user selects the test’s icon (i.e. “Postural Sway” icon), the app guides the user through trial completion via automated instructions.

A Postural Sway (Figure 1A) “trial” is three 10-second upright balance tests, in which the user will stand as still as they can with their hands relaxed at their sides: first, standing with the eyes open and feet apart (EO-FA); second, standing with the eyes open and feet together (EO-FT); finally, standing with eyes closed and feet together (EC-FT).

**Figure 1.**
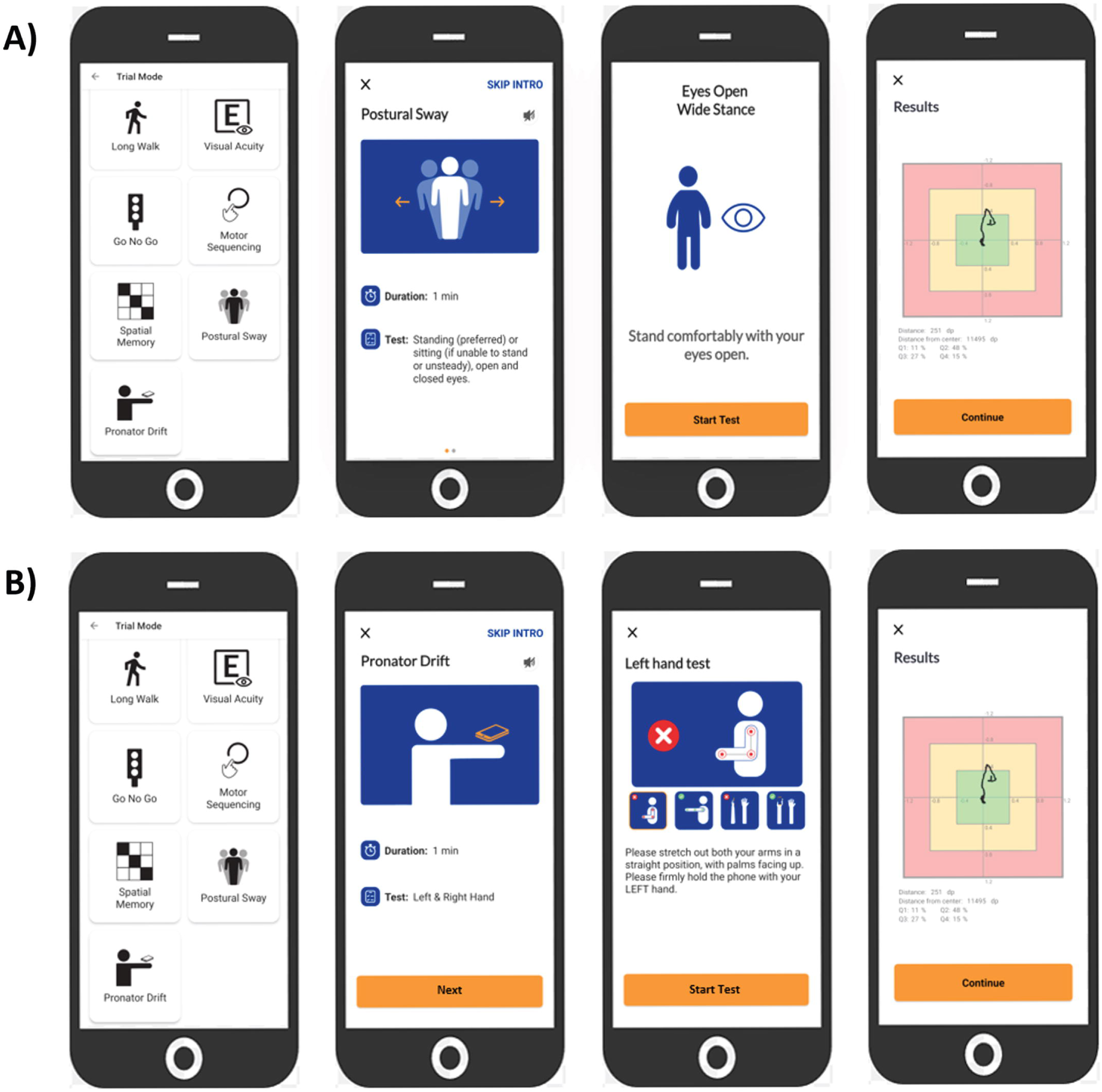
| NeuFun-TS Postural Sway and Pronator Drift user experience walkthrough. **A)** NeuFun-TS Postural Sway app experience. The NeuFun-TS allows users to practice tests, access help tutorials, visualize their history, and submit feedback regarding application bugs. The app currently contains fifteen different tests, which can be accessed in Practice Mode or Trial Mode (shown here). The Postural Sway interface provides both visual and auditory instructions to complete a trial. Upon receiving all instructions, the user may start the trial. The first Postural Sway test evaluates the user’s balance while standing with their feet comfortably apart and their eyes open. After the test is complete, the user’s test movements are plotted to a screen for user feedback, and the user will repeat with their feet together and eyes open and with their feet together and eyes closed. **B)** NeuFun-TS Pronator Drift app overview. The Pronator Drift interface similarly provides visual and verbal guidance for trial completion. Upon receiving all instructions, the user may begin the test. The first Pronator Drift test evaluates the stability of the left hand. After the test is complete, the user’s test movements are plotted to a screen for user feedback. The user will repeat with the right hand.

A Pronator Drift (Figure 1B) “trial” is two 10-second pronator drift tests (one per hand). The user will stand/sit as still as they can with their arms extended forward, palms facing upward, eyes closed, and one hand gripping the phone (strap optional).

A supervising lab member was present during NeuFun-TS testing to answer any questions regarding testing procedure and to ensure subject safety. Each test collects approximately 9.5 seconds of data using the Google Pixel built-in tri-axial accelerometer at a sampling frequency of 50 Hz. Time-series were trimmed to 9-seconds for consistency among comparisons.

### Computing digital biomarkers

Smartphone data was calibrated using an established protocol for accelerometer-based sway analyses in which each test’s 3-Dimensional time series were transformed into one Medio-Lateral (M-L) and one Antero-Posterior (A-P) time series [20]. Additionally, M-L and A-P time series were combined into a single radial acceleration time series, referred to as “Net” acceleration in downstream analyses.

Two time-related and two frequency-related measurements of postural sway and pronator drift were computed based upon review of previous analyses [9–11, 21] (Figure 2).

**Figure 2.**
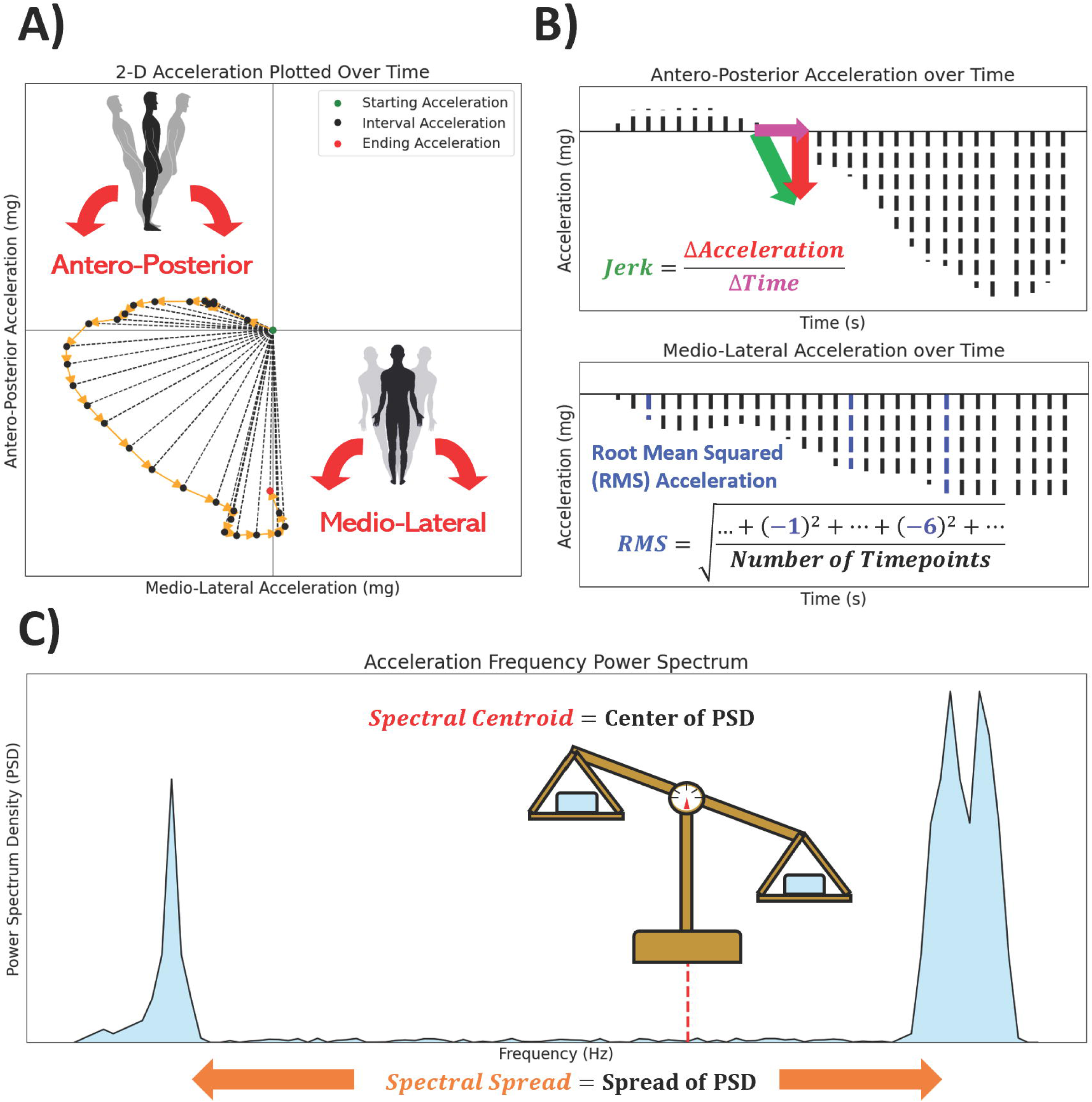
| Visualization of measurements used in the Postural Sway and Pronator Drift analysis. **A)** Manually generated example of 2-Dimensional acceleration data plotted over .6 seconds. Each test’s accelerometry data was converted to Antero-Posterior (“A-P”, front-to-back) and Medio-Lateral (“M-L”, left-to-right) acceleration data. **B)** Display of the M-L component of the 2D acceleration data and provides a visualization of Jerk, which captures the rate of change of acceleration with respect to time, or “sway jerkiness”. **C)** Display of the Antero-Posterior (A-P) component of the 2D acceleration data and a visualization of Root Mean Squared acceleration (RMS), which captures the magnitude of the sway over the course of a test. **D)** Display of frequency-related measurements after transformation of acceleration data. Spectral Centroid captures the central Power Spectrum Density (PSD) of the Power Spectrum, while the Spectral Spread captures extent of deviation of the PSD’s distribution.

Time-related measurements include the following:

1. *Root Mean Square of acceleration (RMS)*, which measures acceleration magnitude. RMS was calculated from the M-L, A-P, and Net acceleration time series as

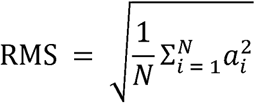 where a_i_ is instantaneous acceleration and N is the number of timepoints.
2. *Sway jerkiness (Jerk)* measures the rate of change in acceleration with respect to time. Jerk was calculated from the M-L and A-P acceleration time series as

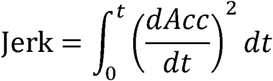 where *t* is time and *Acc* is the M-L or A-P acceleration time series. Net Jerk was calculated using both M-L and A-P acceleration time series (*AccML* and *AccAP*, respectively) as

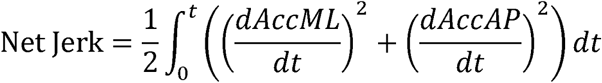 Next, acceleration data were discrete Fourier transformed ^21^ to frequency power spectrums, which captures frequency (Hz) against Power Spectral Density (PSD). From the power spectrums we computed the first two spectral moments μ_1_ and μ_2_, respectively, as our frequency-related measurements:

1. *Spectral centroid (SC)*, which measures the “center of gravity” of the sway frequency (the central PSD-weighted frequency) [21]. SC was calculated from each transformed M-L, A-P, and Net time series as

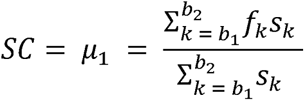 where *b*_1_ and *b*_2_ are the band edges of the frequency samples, *f*_k_ is the k^th^ sample frequency, and *s*_k_ is the k^th^ spectral density.
2. *Spectral spread (SS)*, which measures the variability and dispersion of sway frequency. Following the same notation, SS was calculated from each transformed M-L, A-P, and Net time series as

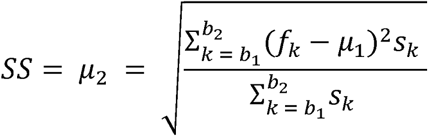

For Postural Sway, each measurement was computed for each stance (EO-FA, EO-FT, EC-FT); differences between testing conditions were evaluated by computing each measurement’s ratio for EC-FT:EO-FT (the Romberg Ratio; measures how removing visual stimulus affects sway) and EO-FT:EO-FA (measures how increasing stance difficulty affects sway).

For Pronator Drift, Net measurements were computed for each hand, which were later relabeled as dominant (“Dom.”) or non-dominant (“Ndom.”) based on subject-identified hand-dominance. Because people with severe disability were holding the phone in the most comfortable manner (strapped sideways or held vertically), we were unable to identify directional movements. We also computed the sums and differences for measurements across dominant and non-dominant hands (“Sum” and “Diff.”), respectively.

Altogether, the different permutations of the four measurements yield 60 Postural Sway digital biomarkers and 16 Pronator Drift digital biomarkers. All biomarkers were log_10_ transformed to improve their distribution normality. Subjects with neurological exam data were randomly split into a training set (80% of subjects) and an independent validation set (20%) before all data analyses (Supplementary Tables 3-6).

### Test-retest reliability and outlier removal

We evaluated inter-day test-retest reliability of derived digital biomarkers by computing the Intraclass Correlation Coefficient (ICC). The ICC compares test-retest variance within subjects with the variance between subjects; high ICC indicates that a biomarker is stable over repeated measures. We computed the ICC (2,1) [22, 23], or the 2-way mixed-effects model, which measures reliability of the first 2 trials of each subject and treats trial number as a rater. Following published guidelines, we used an ICC of 0.5 as a cutoff to remove features with poor test-retest reliability [24].

To assure that downstream analyses are not affected by outliers, we identified outlier biomarker values as those above or below 1.5 times the interquartile range of reliable features.

### Predictive model training and validation

Digitalized tests enable computation of large numbers of digital biomarkers, which can be aggregated into ML models that may outperform individual biomarkers in predicting outcomes. However, ML models can overfit, leading to unrealistically optimistic results. We performed the following steps to mitigate model overfitting:

1. To limit the number of model features, we only included digital biomarkers with ICC>0.5 and correlations with at least one relevant clinical/imaging outcome.
2. To improve model interpretability, we used linear regression models. We selected 3 modeling strategies that differ in level of collinearity stringency to account for redundancy among biomarkers: Ridge regression broadly applies moderate shrinkage to all colinear features (least aggressive); Lasso regression aggressively shrinks redundant features’ coefficients to zero (most aggressive); Elastic Net regression roughly mediates Ridge and Lasso in coefficient optimization.
3. We optimized each model strategy’s parameters through 5-fold cross-validation of the training cohort.
4. For each model strategy, we computed a corresponding component-based model through Principal Component Analysis (PCA).
5. We selected the best model strategy for each outcome by using the coefficient of determination (R^2^).
6. We trained winning models on the complete training cohort and evaluated model performance on the independent validation cohort.
7. We compared model performance in the independent validation cohort with the best single predictor to validate that aggregating biomarkers provides added value over the best single digital biomarker.

We also combined digital biomarkers from both tests and performed steps 1-6 to evaluate how multiple tests model the global disability scales.

## Results

### Digital biomarkers demonstrate moderate inter-day test reproducibility

As detailed in Materials and Methods, we removed biomarkers with ICC below 0.5 as unreliable. From the Postural Sway test, 9 of 60 biomarkers presented moderate reliability (Supplementary Figure 1A). Only two biomarkers, both measuring sway acceleration magnitude, achieved ICC>0.5 for the two open eyes postural sway tests: A-P RMS, which captures acceleration magnitude in antero-posterior directions, and “Net RMS”, which integrates acceleration magnitude from A-P and medio-lateral (M-L) directions. Surprisingly, for the most difficult test (eyes closed, feet together; EC-FT), in addition to RMS biomarkers, M-L Jerk and Net Jerk achieved ICC>0.5. In fact, ICC for M-L RMS EC-FT was much higher (i.e., 0.695) than that of A-P RMS (i.e., 0.535-0.566 for all 3 stances). This suggests that vision controls postural sway in M-L directions much more effectively than in A-P directions and eliminating this visual correction increases test sensitivity.

For Pronator Drift (Supplementary Figure 1B), 5 digital biomarkers achieved ICC>0.5. Again, RMS exhibited stronger reproducibility than acceleration jerk, but only for the dominant hand.

### Postural sway digital biomarkers correlate with age

Previous postural sway research has identified in healthy subjects a positive correlation between accelerometry-derived measurements and age [25]. Some research has also indicated correlations are present between height, weight, and sex [26, 27]. For each diagnosis type with at least 10 subjects, we computed correlations between the reliable features and age, sex, height, and weight. Net RMS EC-FT demonstrated the strongest correlation in HV, and was the only biomarker to correlate with age in all cohorts with 10 or more subjects (Figure 3A; Supplementary Tables 11-14). Net RMS EC-FT explained 67% and 57% of variance in HV and NIND cohorts, respectively (p<0.001), with almost identical slopes and intercepts. This indicates that Net RMS EC-FT measures age-related increase in postural sway. Other postural sway biomarkers correlated with age and one pronator drift biomarker correlated with sex, although sex distribution was limited (Supplementary Figure 2).

**Figure 3.**
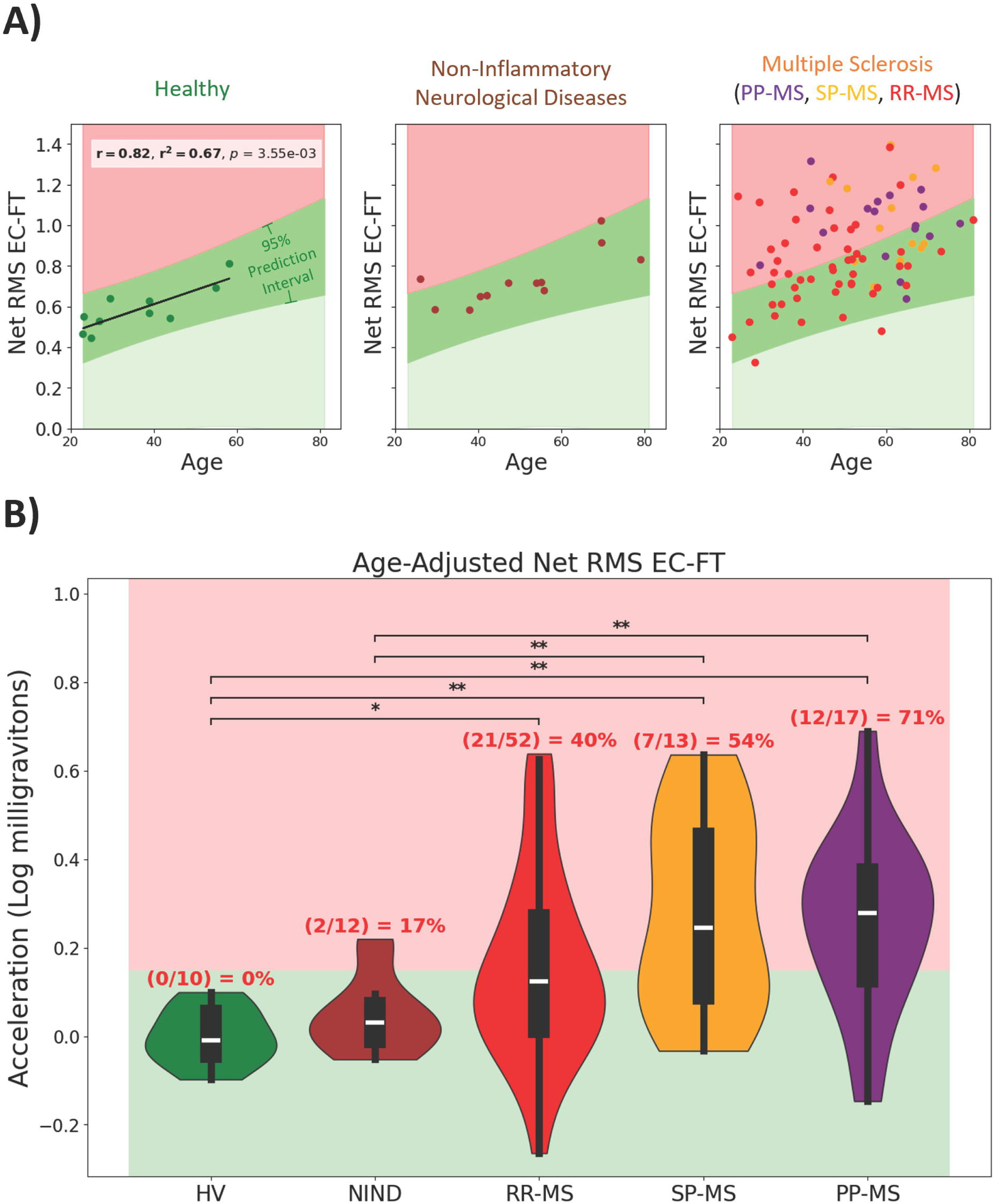
| Age exhibits differentiable relationship among cohorts’ sway amplitude measurements. **A)** Net RMS EC-FT (Net sway amplitude for Eyes Closed and Feet Together) significantly correlated with age in every diagnosis cohort with at least 10 subjects (HV, NIND, and MS). Pearson’s (r) correlations and their p-values (p) were adjusted using the Benjamini-Hochberg False Discovery Rate adjustment with alpha = .05. **B)** Age-adjusted Net RMS EC-FT prediction interval identifies abnormal digital biomarker results corresponding to MS-related disability. The 95% prediction interval was calculated from the Healthy Volunteer (HV) cohort and was used to identify abnormal Net RMS EC-FT in subjects with Multiple Sclerosis (MS), Non-Inflammatory Neurological Diseases (NIND), Relapsing-Remitting MS (RR-MS), Secondary-Progressive MS (SP-MS), and Primary-Progressive MS (PP-MS). The Wilcoxon Rank-Sum nonparametric test of difference was performed pairwise among all diagnostic groups using Benjamini Hochberg False Discovery Rate corrections (alpha = 0.05) to adjust p-values; * corresponds to significant differences with p-value <= 0.05, ** corresponds to p-value <= 0.01.

After subtracting the effect of natural aging (i.e., using Net RMS EC-FT HV age residuals), we observed that Net RMS EC-FT differentiates MS patients from HV and NIND controls (Figure 3B). 17% of NIND subjects, 40% of RRMS, 54% of SPMS and 71% of PPMS patients measured Net RMS EC-FT beyond the effect of natural aging. Thus, Net RMS EC-FT also differentiates MS-related increase in postural sway from natural aging.

### Postural sway digital biomarkers correlate more strongly with relevant clinical and imaging outcomes than pronator drift biomarkers

Next, we aimed to select the most clinically relevant digital biomarkers for modeling. Because biomarker-derived models must be blindly tested in the independent validation cohort, we only used training cohort data for biomarker selection (Supplementary Tables 15-18).

We computed Pearson correlations between reliable Postural Sway and Pronator Drift features and 3 global neurological disability scales (EDSS: ordinal, from 0-10; CombiWISE: ML-learning derived, continuous, from 0-100; NeurEx^TM^: derived from NeurEx^TM^ App, continuous linear scale, theoretical maximum of 1347). Because postural sway and pronator drift tests measure subdomains of neurological examination, we also evaluated biomarker correlations with relevant subpanels, detailed in Materials in Methods, of the aforementioned global scales (Figure 4).

**Figure 4.**
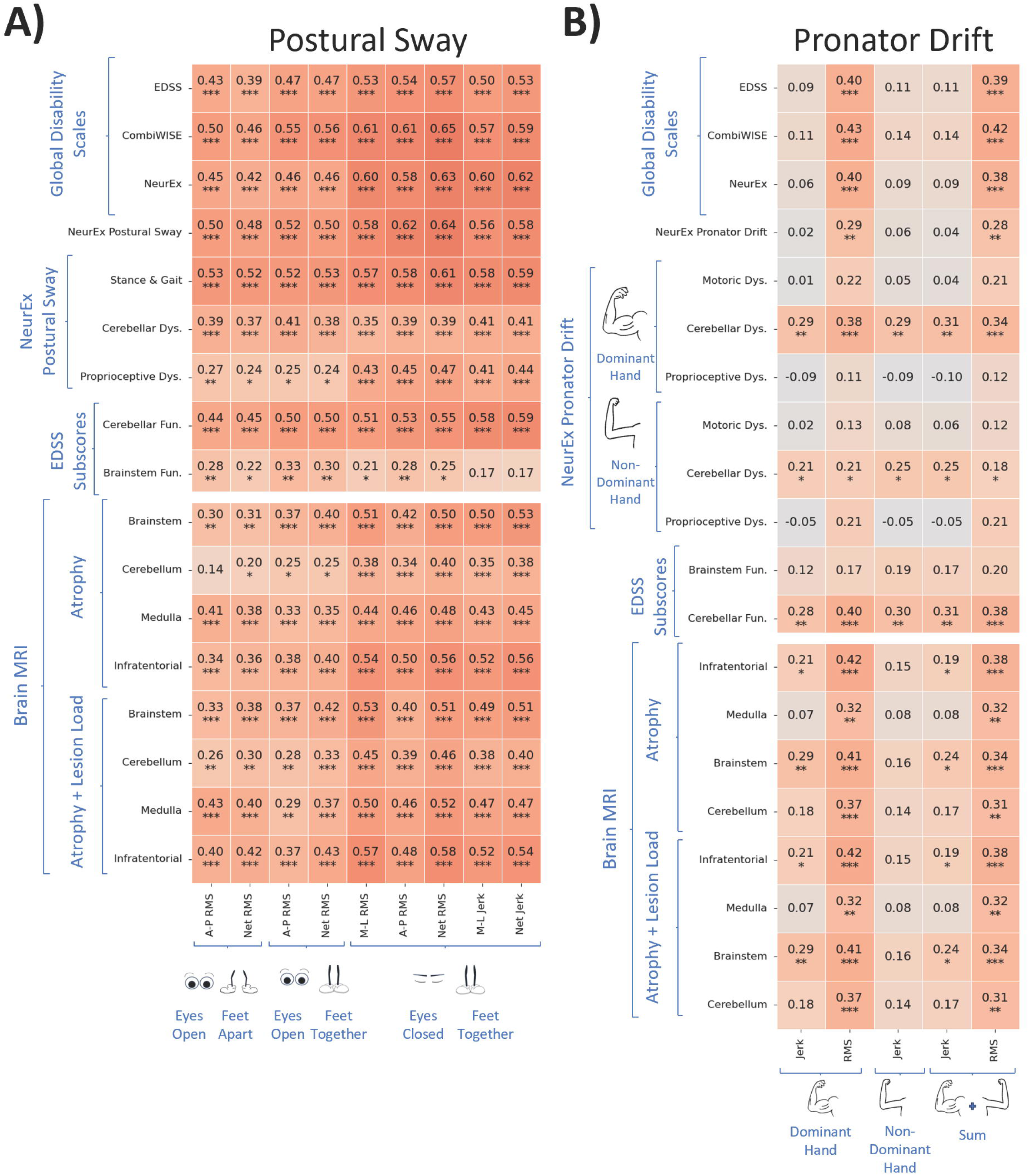
| Digital biomarkers exhibit low to moderately strong Pearson correlations with Clinical Scores and MRI Scores. **A)** Postural Sway features exhibit significant, moderate Pearson correlations. **B)** Pronator Drift features exhibit significant, low Pearson correlations with Clinical Scores and MRI Scores. Correlation p-values were adjusted using the Benjamini-Hochberg False Discovery Rate adjustment with alpha = .05. RMS refers to root mean squared acceleration, which captures sway amplitude; Jerk refers to the rate of change of acceleration, which captures sway jerkiness. A-P refers to antero-posterior movement, which captures forward and backward sway; M-L refers to medio-lateral movement, which captures side-to-sway sway; Net combines M-L and A-P acceleration data to capture overall acceleration. “Dys.” is short for “Dysfunction”, “Fun.” is short for “Function”. Benjamini Hochberg False Discovery Rate corrections (alpha = 0.05) were used to adjust p-values; * corresponds to significant differences with p-value <= 0.05, ** corresponds to p-value <= 0.01.

All postural sway digital biomarkers correlated moderately strongly (r>0.5, p<0.001) with at least one clinical outcome. We observed a hierarchy in Pearson’s correlations with global disability scales: correlations were weakest with EDSS (r=0.39-0.57, p<0.001), stronger with NeurEx (r=0.42-0.63, p<0.001) and strongest with CombiWISE (r=0.46-0.65, p<0.001). As expected, most digital biomarkers correlated comparatively stronger with subpanels of global disability scales, such as the stance and gait subpanel (r=0.52-0.61, p<0.001) or NeurEx^TM^ Postural Sway (r=0.48-0.64, p<0.001). EC-FT biomarkers correlated stronger with the proprioception subpanel (r=0.41-0.47, p<0.001) than eyes open (EO-FA, EO-FT) biomarkers (r=0.24-0.27, p<0.05). This indicates that vision can largely compensate for the effect of decreased proprioception.

Additionally, all postural sway digital biomarkers correlated moderately with semi-quantitative imaging outcomes in the infratentorial compartment (Figure 4A), although these correlations were slightly weaker (r=0.34-0.58) compared to correlations with clinical outcomes (r=0.35-0.65). Generally, all 3 infratentorial sites (brainstem, cerebellum and medulla/upper cervical spine) contributed to these correlations. The infratentorial scores that integrate atrophy of all infratentorial anatomical regions correlated stronger with digital biomarkers than atrophy of each individual CNS anatomical site (r=0.34-0.58 vs r=0.14-0.51). Likewise, scores that aggregated both lesion load and atrophy correlated stronger (r=0.26-0.58) compared to semiquantitative atrophy scores only (r=0.14-0.56).

Net RMS EC-FT outperformed all digital biomarkers in correlations with clinical and imaging outcomes with the exception of correlations with cerebellar subpanels that correlated strongest with A-P RMS EO-FT stance, consistent with clinical knowledge that vision does not compensate for cerebellar dysfunction in postural sway.

Pronator Drift biomarkers exhibited comparatively weaker correlations overall. Within pronator drift biomarkers, Jerk correlated weaker than RMS (r=-0.05 to 0.29, p<0.01 vs r =0.11-0.43, p<0.001). Surprisingly, biomarkers did not correlate with motoric and proprioceptive subpanels but only with cerebellar subpanels (up to r = 0.43, p<0.001). Because NeurEx^TM^ Pronator Drift integrates motoric, proprioceptive and cerebellar dysfunction, the subpanel also did not correlate with any digital biomarker. The best biomarker from Pronator Drift was Net RMS Dom., which correlated with all global disability scales and demonstrated the same hierarchy we observed for postural sway biomarkers: EDSS<NeurEx<CombiWISE (r=0.40-0.43, p<0.001). RMS Dom. also correlated with all imaging outcomes (r=0.32-0.42, p<0.001).

### Development and validation of digital biomarker models

We sought to predict 3 global scales of neurological disability (EDSS, CombiWISE, NeurEx^TM^) and 2 test-specific scales (NeurEx^TM^ Postural Sway, NeurEx^TM^ Pronator Drift) using model strategies that accounted for multicollinearity among digital biomarkers (Supplementary Figure 3). However, because pronator drift digital biomarkers did not correlate with NeurEx^TM^ Pronator Drift, we instead used them to model NeurEx^TM^ Cerebellar Dysfunction Dom., which these biomarkers correlated with (Figure 4B), as a sensitivity analysis.

After optimizing model parameters and ensuring that models performed comparably to principal component models (Supplementary Figures 4-6), we selected the best model type (Ridge, Lasso, or Elastic Net) using the coefficient of determination (R^2^). We trained these models on the complete training cohort (Supplementary Figures 7-9) and tested them in the independent validation cohort (Figure 5; Supplementary Figures 10-11). Pearson’s correlation (r), the coefficient of determination, and the concordance correlation coefficient (CCC) were used as model performance metrics.

**Figure 5.**
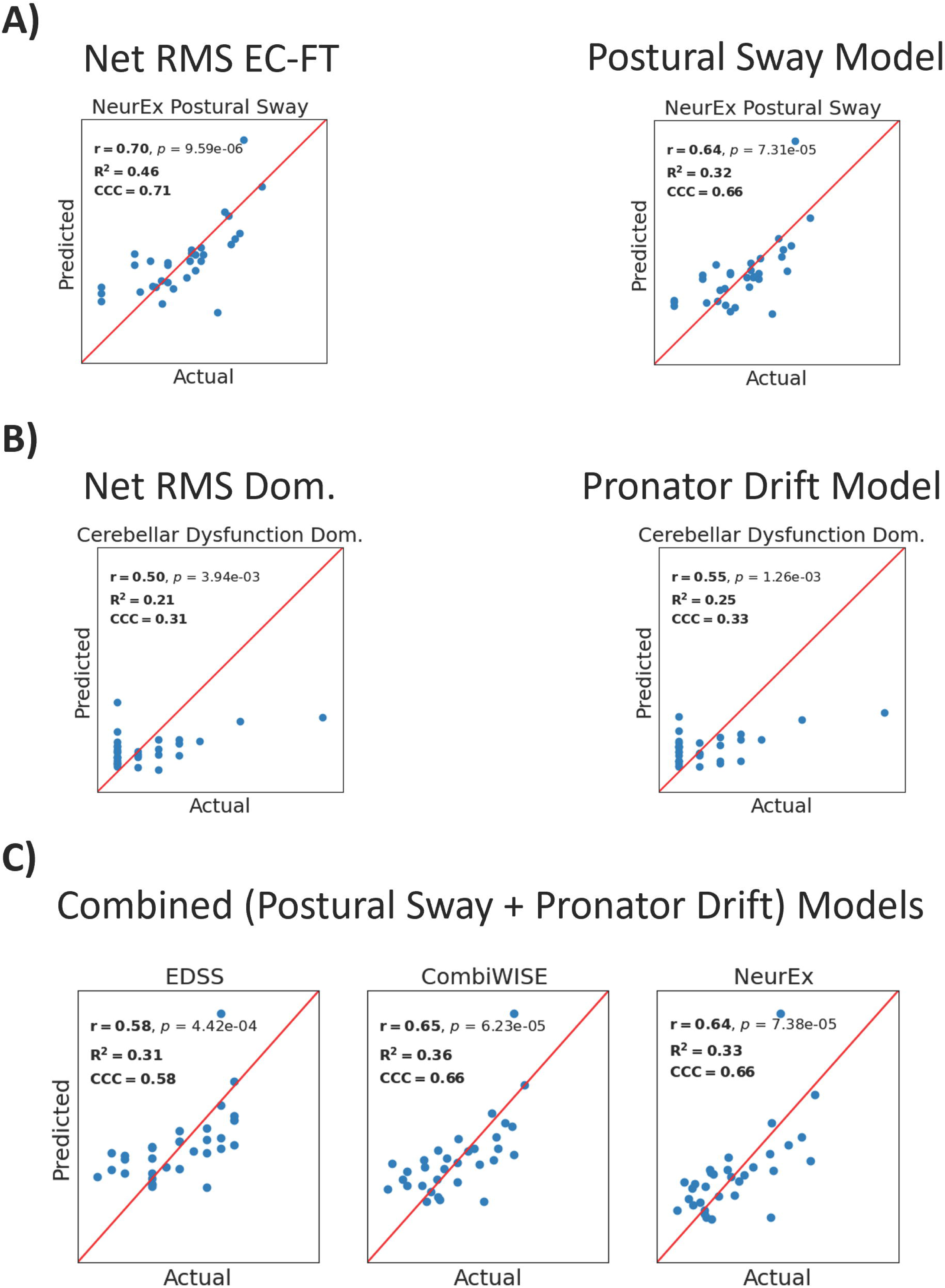
| Performance metrics of single best predictors and best models in independent validation cohort. Pearson’s (r) correlation, the coefficient of determination (R^2^), and the concordance correlation coefficient (CCC) were used as metrics of model success. **A)** The best single predictor for the NeurEx^TM^ Postural Sway subpanel was Net RMS EC-FT, which achieves higher prediction and correlation coefficients than the best model assembled from all postural sway biomarkers. **B)** The best model assembled from all pronator drift biomarkers outperformed the best single predictor for pronator drift Net RMS Dom. in predicting the NeurEx^TM^ Cerebellar Dysfunction subpanel for the dominant hand. **C)** Combining biomarkers from the postural sway and pronator drift tests yields models that predict global scales of disability better than the postural sway or pronator drift test alone. RMS refers to root mean squared acceleration, which captures sway amplitude. Net combines Medio-Lateral and Antero-Posterior acceleration data to capture overall acceleration. “Dom.” is short for “Dominant hand”.

First, we evaluated the ability of test-specific models (i.e., models derived from Postural Sway biomarkers) to predict test-specific outcomes (i.e., NeurEx^TM^ Postural Sway); we compared these models with the best single predictors for each model outcome. Surprisingly, for predicting NeurEx^TM^ Postural Sway, we found that the best single predictor achieved greater model performance metrics than the best Postural Sway model (r=.70, R^2^=.46, CCC=.71 versus r=.64, R^2^=.38, CCC=.68, respectively; Figure 5A). For Pronator Drift modeling we found that the best Pronator Drift model outperformed the best single predictor in predicting cerebellar dysfunction in the validation cohort (r=.55, R^2^=.25, CCC=.33 versus r=.50, R^2^=.21, CCC=.31, respectively; Figure 5B).

Next, we evaluated whether models that combine Postural Sway and Pronator Drift biomarkers outperform the best test-specific models in predicting global disability scales. Indeed, combined models achieved the highest performance metrics for EDSS, CombiWISE, and NeurEx^TM^ (r=.58-65, R^2^=.31-.36, CCC=.58-.66), indicating that accurately measuring global disability requires integrating biomarkers from multiple functional tests (Figure 5C; Supplementary Figures 10-11).

## Discussion

### Key Findings

Analysis of the simple, 1-minute NeuFun-TS Postural Sway test yielded a digital biomarker (Net RMS EC-FT) with moderately strong test-retest reproducibility, meaningful correlations with age (even in HV), and predictive power for disability scores. This biomarker’s correlation with age aligns with previous studies that suggest age-related degeneration in proprioception and delayed motoric integration contributes to increased postural instability in older adults [28, 29]. Our findings further previous research by uncovering how healthy subjects and subjects with non-inflammatory neurological diseases exhibit a nearly identical progression of disability with age, while subjects with progressive forms of MS (Secondary and Primary Progressive) followed steeper slopes. This identified MS-specific contributions to postural sway dysfunction. Notably, Net RMS EC-FT, which had the highest test-retest reliability, outperformed postural sway models that incorporated some less reliable biomarkers; establishing reliability of digital biomarkers is essential to functional test analyses.

Moreover, while the NeuFun-TS Pronator Drift test also yielded moderately reliable digital biomarkers, the biomarkers correlated weaker with clinical outcomes. Nevertheless, these weaker biomarkers provided additional predictive value when we combined them with postural sway biomarkers to predict global scales of neurological disability. This highlights the importance of having functional tests that cover different (ideally all) neurological subsystems in the NeuFun-TS in order to capture development of global neurological disability.

### Future directions

While the goal of NeuFun-TS is measuring all neurological subsystems, the time required to perform all tests determines NeuFun-TS usability and, consequently, testing compliance. Therefore, NeuFun-TS must balance test accuracy (may require longer testing times) and usability (requires short testing). With this in mind, we will remove Pronator Drift from NeuFun-TS, because the test is only sensitive for cerebellar dysfunction, which is already measured with greater accuracy in other NeuFun-TS test [17]. Likewise, Postural Sway EO-FA position did not yield any non-redundant digital biomarkers. Thus, we will explore whether abandoning this position and increasing time for EO-FT, which is sensitive to cerebellar dysfunction, and EC-FT, which provides the best digital biomarker of Postural Sway, may enhance reliability of these clinically meaningful biomarkers.

### Potential limitations

One limitation of the postural sway test, as previously mentioned, was that the distance between the feet for the feet apart stance was not controlled between subjects; however, because this position did not provide useful biomarkers we will be removing it from the NeuFun-TS. One potential limitation of the pronator drift test was that orientation of the smartphone was not controlled, which prevented directional movements from being captured. However, signal interference from hand tremors and the inability of the digital biomarkers to discern pronation/drift in most subjects means that accelerometers would likely fail to capture directional movements regardless.

## Conclusion

The user-friendly, 1-minute NeuFun-TS Postural Sway test exhibits meaningful correlations with age and clinician scores reflecting balance. Assembling models from different NeuFun-TS tests yields models better able to predict clinical outcomes.

## Data Availability

Data and code are available at

https://github.com/mcalcagninih/PosturalSway_PronatorDrift_Code

## Abbreviations

CCC: Concordance correlation coefficient, which measures absolute agreement between two variables
CIS: Clinically isolated syndrome
Dom.: dominant-hand results from the pronator drift test
HV: healthy volunteers
ICC: Intraclass correlation coefficient, which measures test-retest reproducibility/reliability
EC-FT: “Eyes closed feet together”; refers to digital biomarkers from the first stance in the postural sway test in which the subject stands with eyes closed and feet together
EO-FA: “Eyes open feet apart”; refers to digital biomarkers from the first stance in the postural sway test in which the subject stands with eyes open and feet apart
EO-FT: “Eyes open feet together”; refers to digital biomarkers from the second stance in the postural sway test in which the subject stands with eyes open and feet together
EN: Elastic net, a regularized regression modeling strategy that combines Lasso and Ridge regression penalization effects
MRI: Magnetic resonance imaging
MS: Multiple sclerosis
OIND: Other inflammatory neurological diseases
NIAID: National Institute of Allergy and Infectious Diseases
NIH: National Institutes of Health
NIND: Non-inflammatory neurological diseases
NeuFun-TS: Neurological Function Tests Suite, a smartphone application containing sixteen different functional tests (including the NeuFun-TS Postural Sway and NeuFun-TS Pronator Drift tests) used to evaluate neurological disability in different domains
PCA: Principle component analysis
PP-MS: Primary Progressive Multiple Sclerosis
RIS: Radiologically isolated syndrome
RMS: Root Mean Squared; refers to the acceleration magnitude, an accelerometry-derived measurement, which was computed by squaring, averaging, and square-root transforming accelerometer time series data
RR-MS: Relapsing-remitting multiple sclerosis
SP-MS: Secondary progressive multiple sclerosis

## Declarations

### Ethics Approval and Consent to Participate

This study was reviewed and approved by National Institute of Allergy and Infectious Diseases (NIAID) scientific review and the National Institutes of Health (NIH) Institutional Review Board. All participants provided written or digital informed consent to participate in this study.

### Consent for Publication

All participants provided written or digital informed consent for publication.

### Availability of Data and Materials

Data and code are available at https://github.com/mcalcagninih/PosturalSway_PronatorDrift_Code. All data analyses were performed using Python [22, 23, 30].

## Acknowledgements

We would like to thank the Neuroimmunological Diseases Section (NDS) clinical team for patient scheduling and expert patient care. We would also like to thank all previous post-baccalaureate researchers who assisted in clinical testing. Finally, we would like to thank all patients, caregivers, and healthy volunteers, without whom this research would not be possible.

## Author Contributions

M.C. guided research subjects through smartphone testing, maintained cloud database and smartphone updates, curated and preprocessed smartphone data, performed data analyses, designed and prepared figures and tables, and drafted the main manuscript text. P.K. curated clinical score and imaging data, guided data analyses, and guided figure and table design. B.B. conceptualized and designed the study, guided data analyses, guided figure and table design, performed neurological examinations, and edited the main manuscript text. All authors reviewed the manuscript.

