## Supplementary Material legends for "Smartphone Postural Sway and Pronator Drift tests as Measures of Neurological Disability"

**Supplementary Figure Legends**

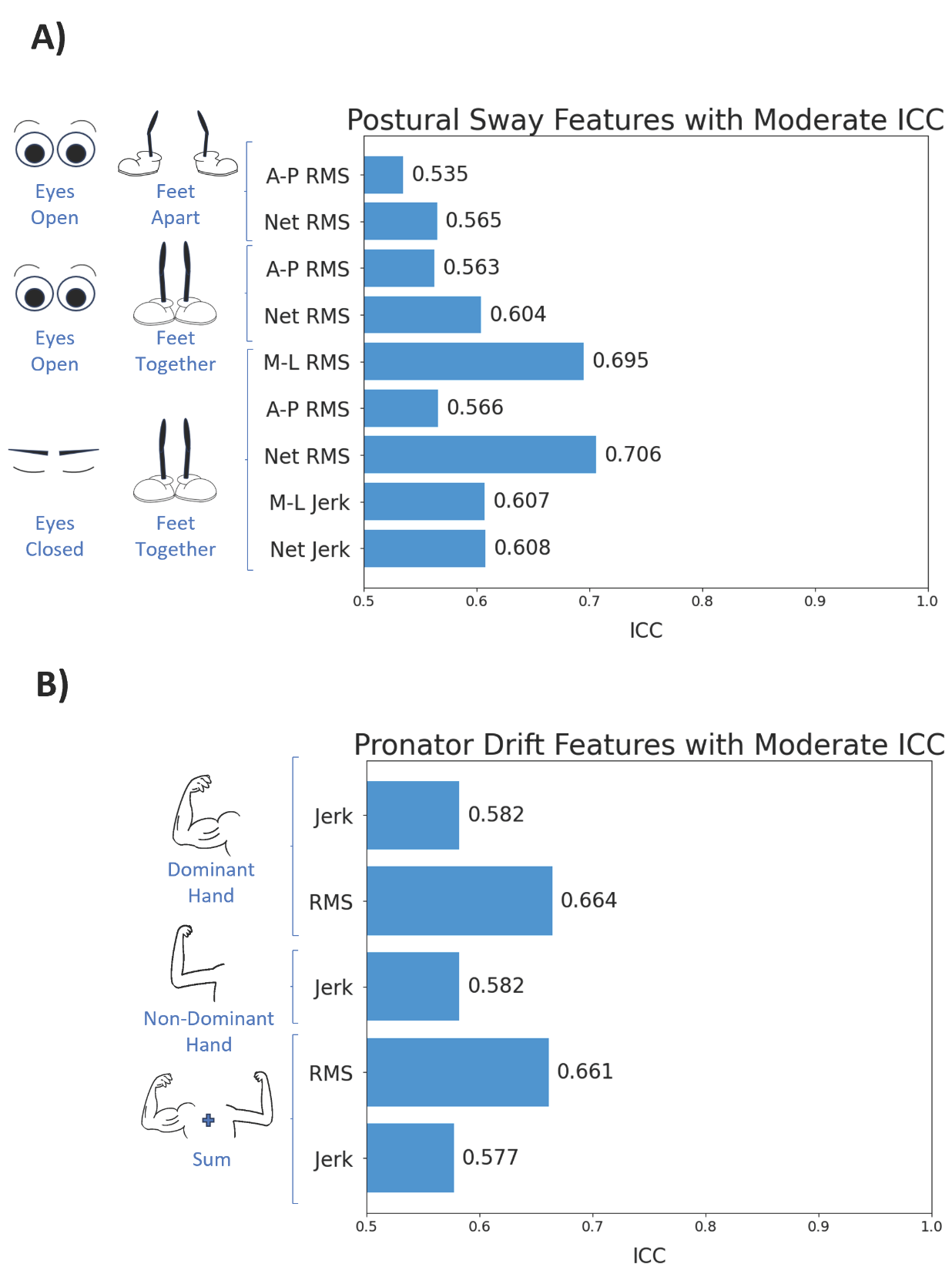

Supplementary Figure 1 | The intraclass correlation coefficient (ICC) for test-retest reliability identified reliable features from the Postural Sway and Pronator Drift tests. A) 9 Postural Sway features (digital biomarkers) exhibited moderate reliability (ICC at least .5), spanning all three testing conditions. B) 5 Pronator Drift test features had moderate reliability from both dominant and non-dominant hands. All p-values were adjusted using the Benjamini-Hochberg False Discovery Adjustment with alpha = .05.

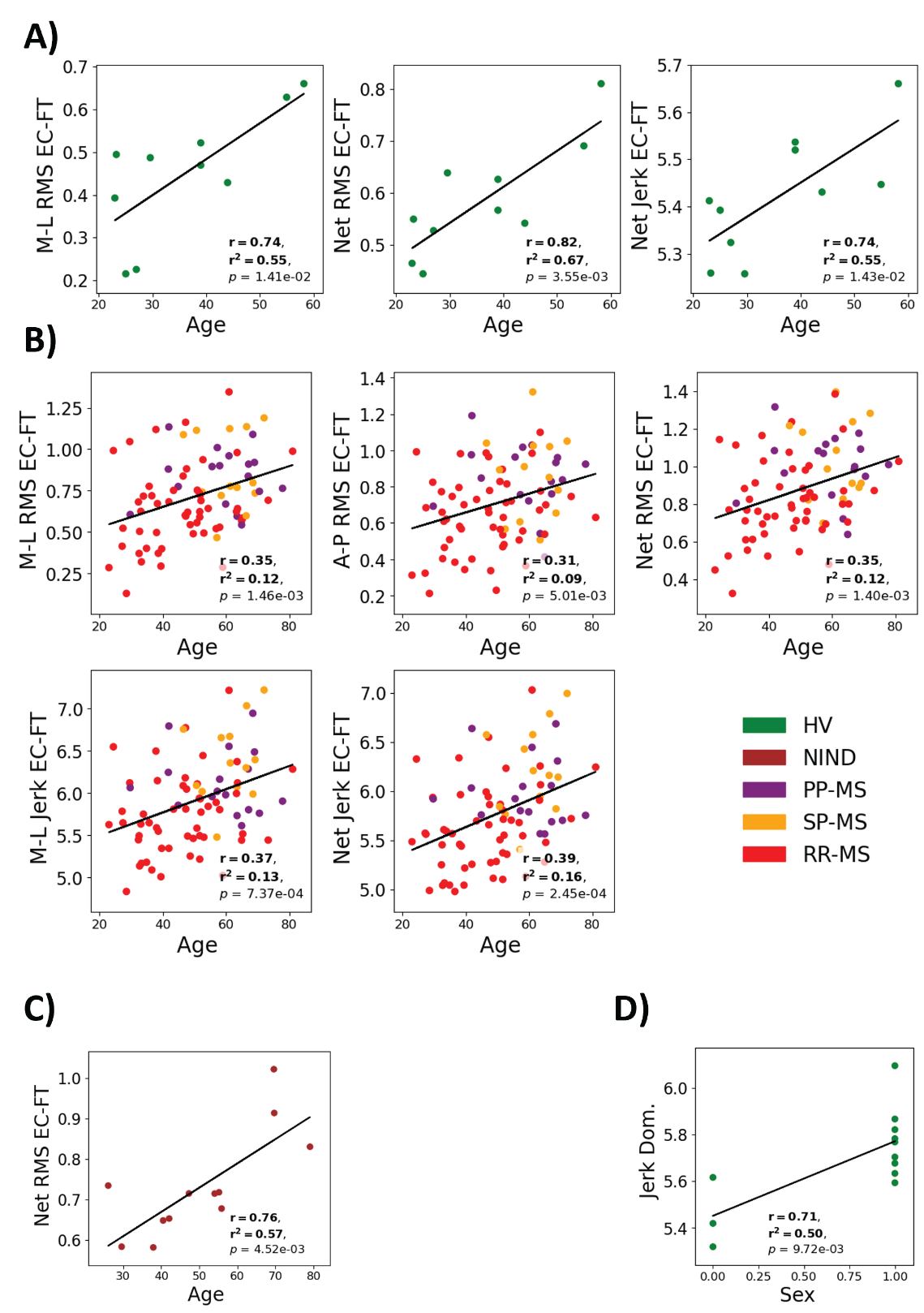

Supplementary Figure 2 | Significant Pearson correlations between digital biomarkers and age and sex. A) Postural Sway biomarkers correlate with age in the healthy volunteer (HV) cohort. B) A Postural Sway biomarker correlates with age in the cohort of subjects with Non-Inflammatory Neurological Diseases (NIND). C) Postural Sway biomarkers correlate with age in the cohort of subjects with Multiple Sclerosis (MS); this cohort includes subjects with Relapsing-Remitting MS (RR-MS), Primary Progressive MS (PP-MS), and Secondary Progressive MS (SP-MS). D) A Pronator Drift biomarker correlates with sex in the HV cohort. * indicates p-value < .05, ** indicates p-value < .01, *** indicates p-value < .001. All p-values were adjusted using the Benjamini-Hochberg False Discovery adjustment with alpha = .05.

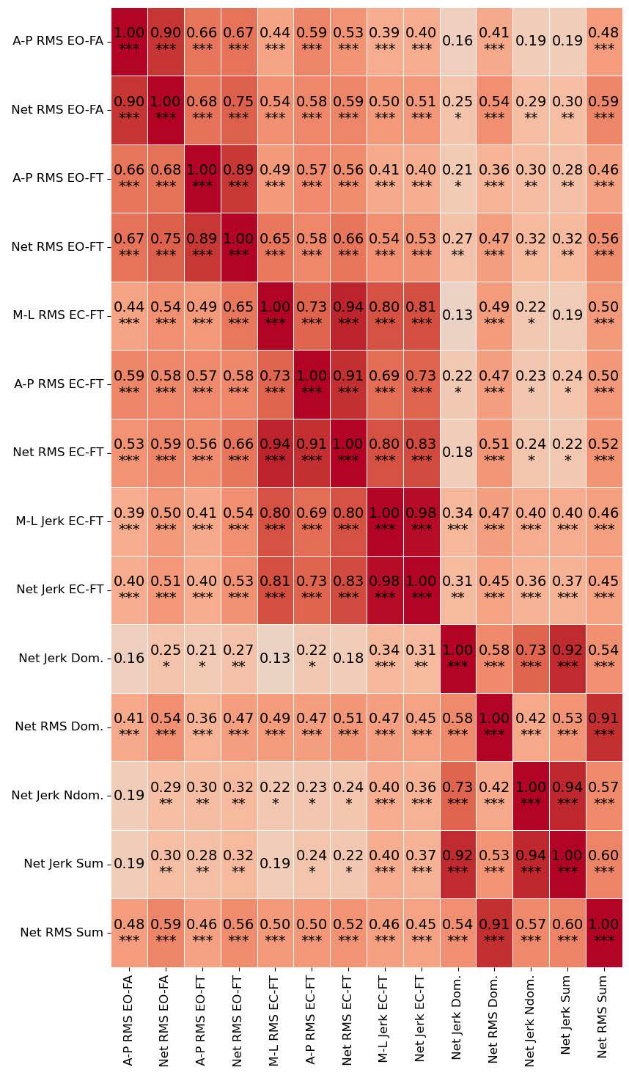

Supplementary Figure 3 | Pearson correlations among all analyzed model features (digital biomarkers) from both postural sway and pronator drift tests. * indicates p-value < .05, ** indicates p-value < .01, *** indicates p-value < .001. All p-values were adjusted using the Benjamini-Hochberg False Discovery adjustment with alpha = .05.

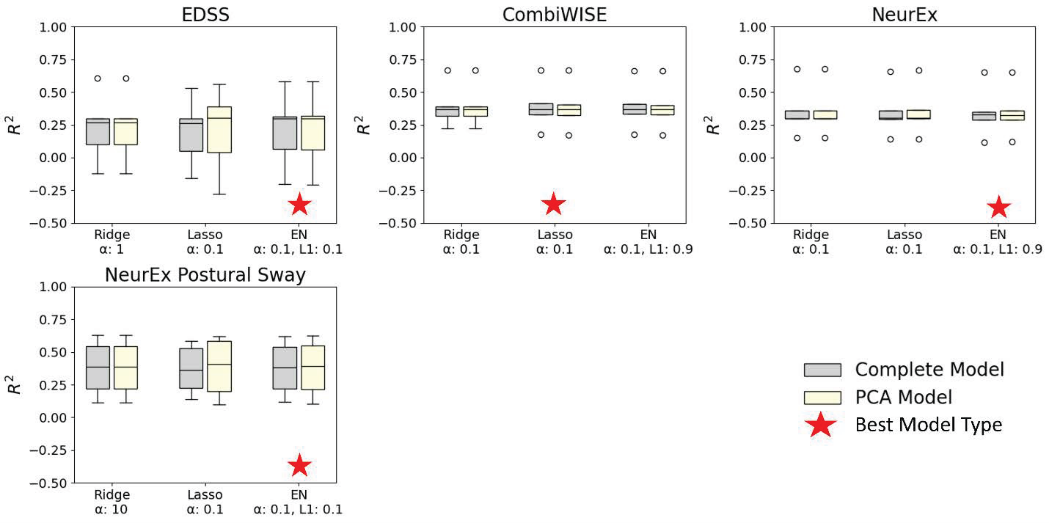

Supplementary Figure 4 | Cross-validation results for models derived from Postural Sway digital biomarkers. Complete models (models composed of linear combinations of all biomarkers) demonstrated comparable results with principal component models (PCA). The model strategies and their corresponding parameters are listed on the x-axes; the coefficient of determination (R^2^) is on the y-axis.

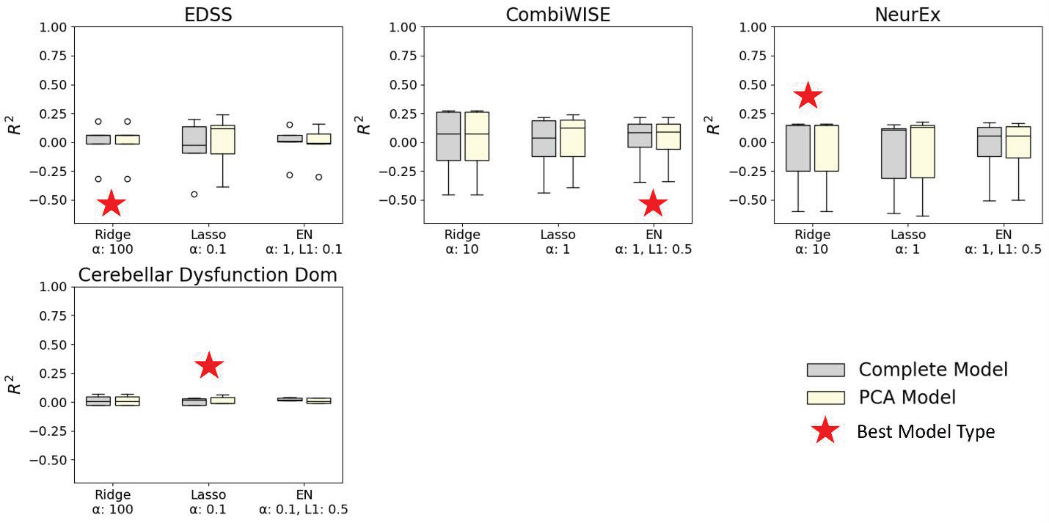

Supplementary Figure 5 | Cross-validation results for models derived from Pronator Drift digital biomarkers. Complete models (models composed of linear combinations of all biomarkers) demonstrated comparable results with principal component models (PCA). The model strategies and their corresponding parameters are listed on the x-axes; the coefficient of determination (R^2^) is on the y-axis.

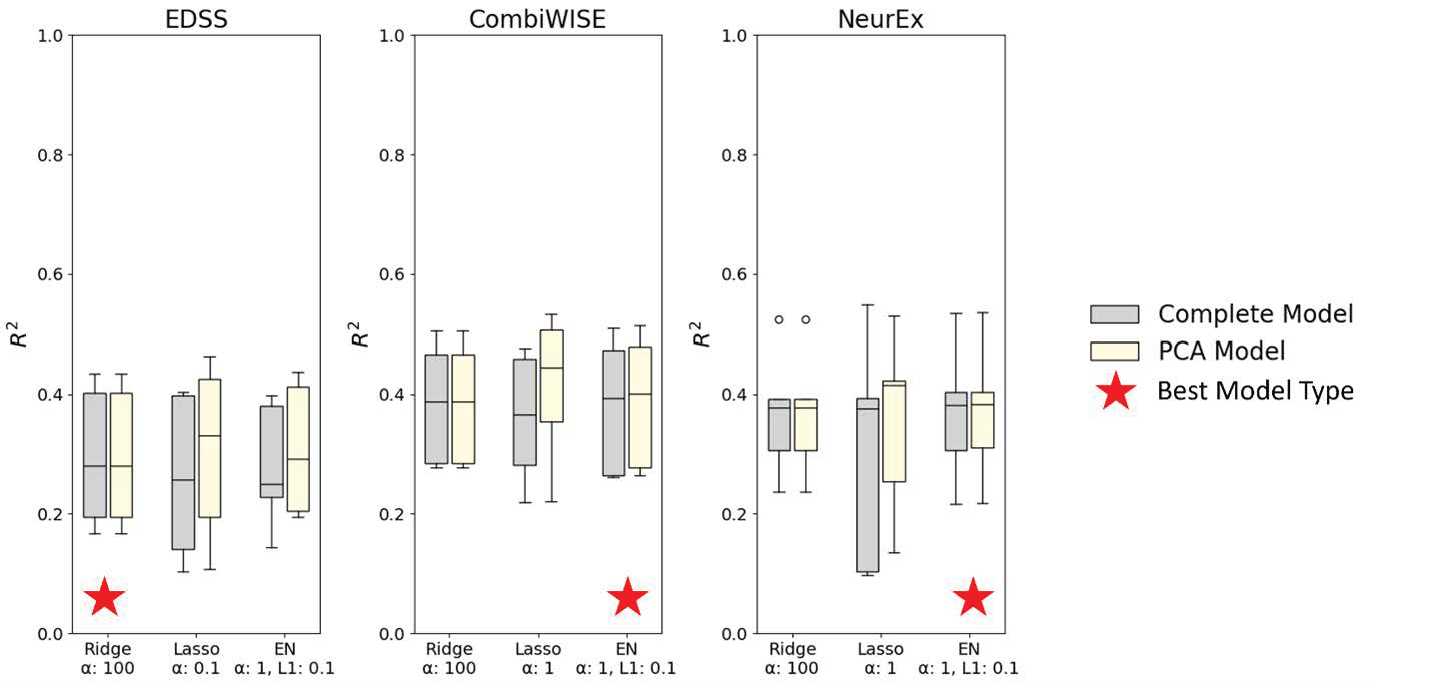

Supplementary Figure 6 | Cross-validation results for models derived from Postural Sway and Pronator Drift digital biomarkers. Complete models (models composed of linear combinations of all biomarkers) demonstrated comparable results with principal component models (PCA). The model strategies and their corresponding parameters are listed on the x-axes; the coefficient of determination (R^2^) is on the y-axis.

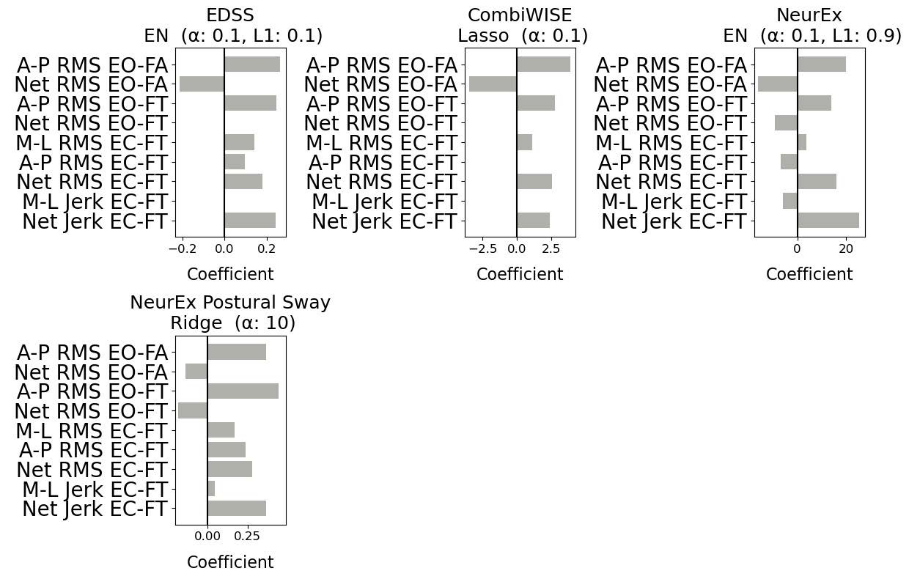

Supplementary Figure 7 | Coefficients for all winning models derived from Postural Sway digital biomarkers. The model outcome being predicted (i.e. EDSS) is listed above the model strategy (Ridge, Lasso, or Elastic-Net).

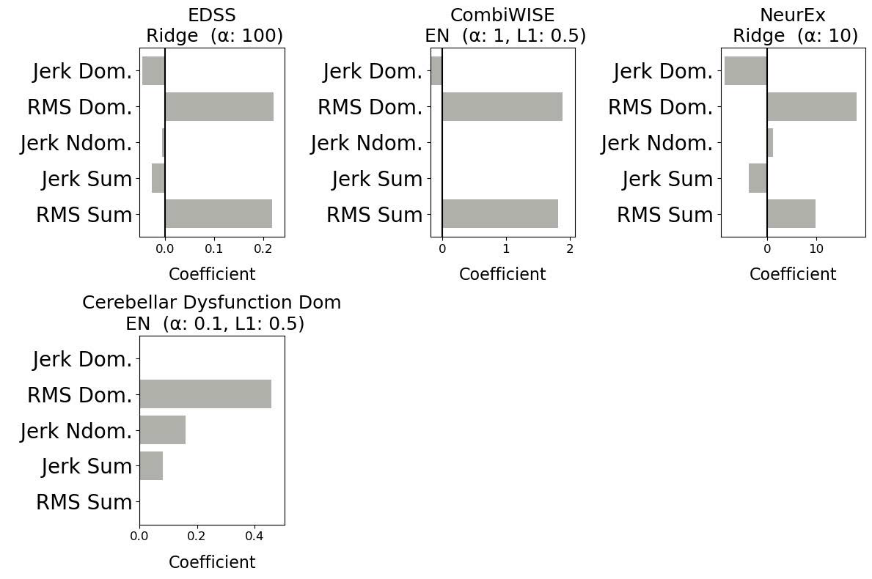

Supplementary Figure 8 | Coefficients for all winning models derived from Pronator Drift digital biomarkers. The model outcome being predicted (i.e. EDSS) is listed above the model strategy (Ridge, Lasso, or Elastic-Net).

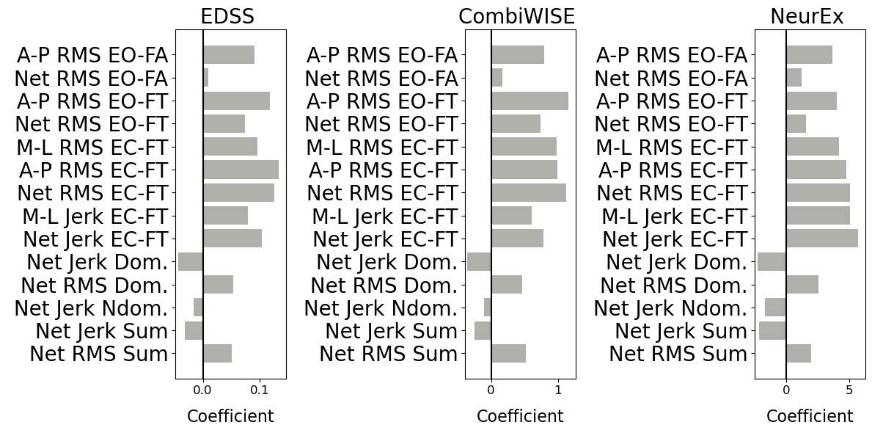

Supplementary Figure 9 | Coefficients for all winning models derived from Postural Sway and Pronator Drift digital biomarkers. The model outcome being predicted (i.e. EDSS) is listed above the model strategy (Ridge, Lasso, or Elastic-Net).

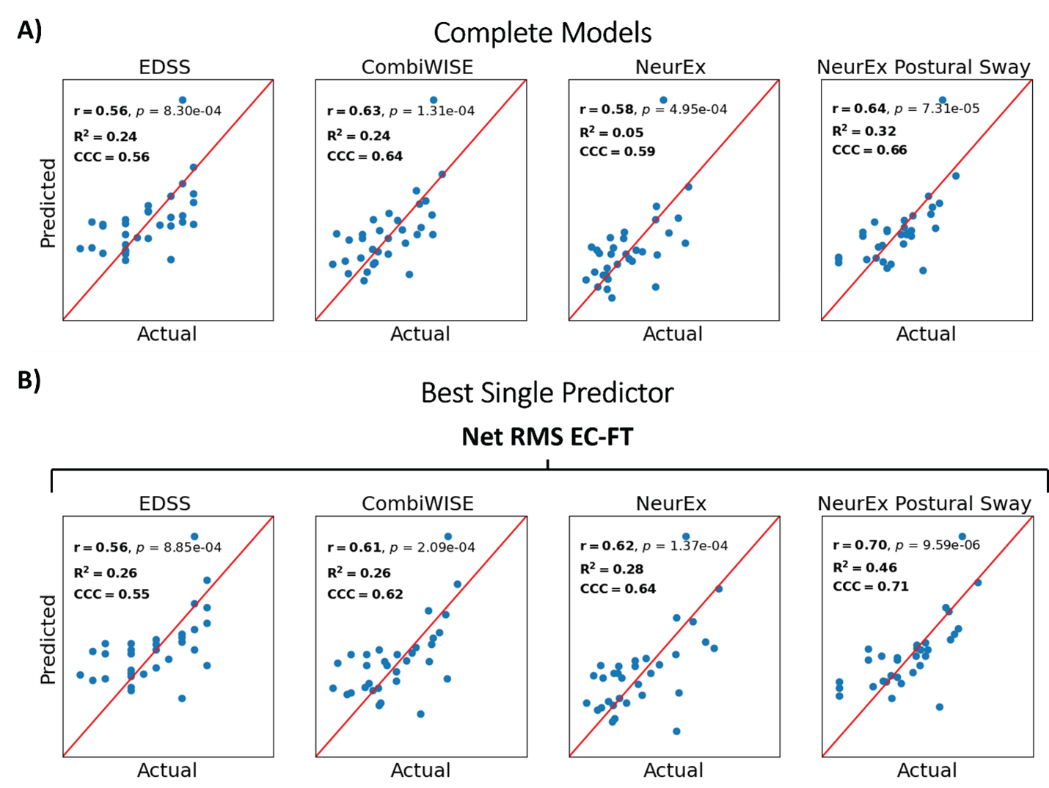

Supplementary Figure 10 | Independent validation results for predictive models cohort compared to the best single predictor for each model outcome (EDSS, CombiWISE, NeurEx^TM^, NeurEx^TM^ Postural Sway). using Postural Sway digital biomarkers. Pearson’s r, the coefficient of determination (R^2^), and the concordance correlation coefficient (CCC) were used to evaluate models’ predictive strength for their respective outcomes.

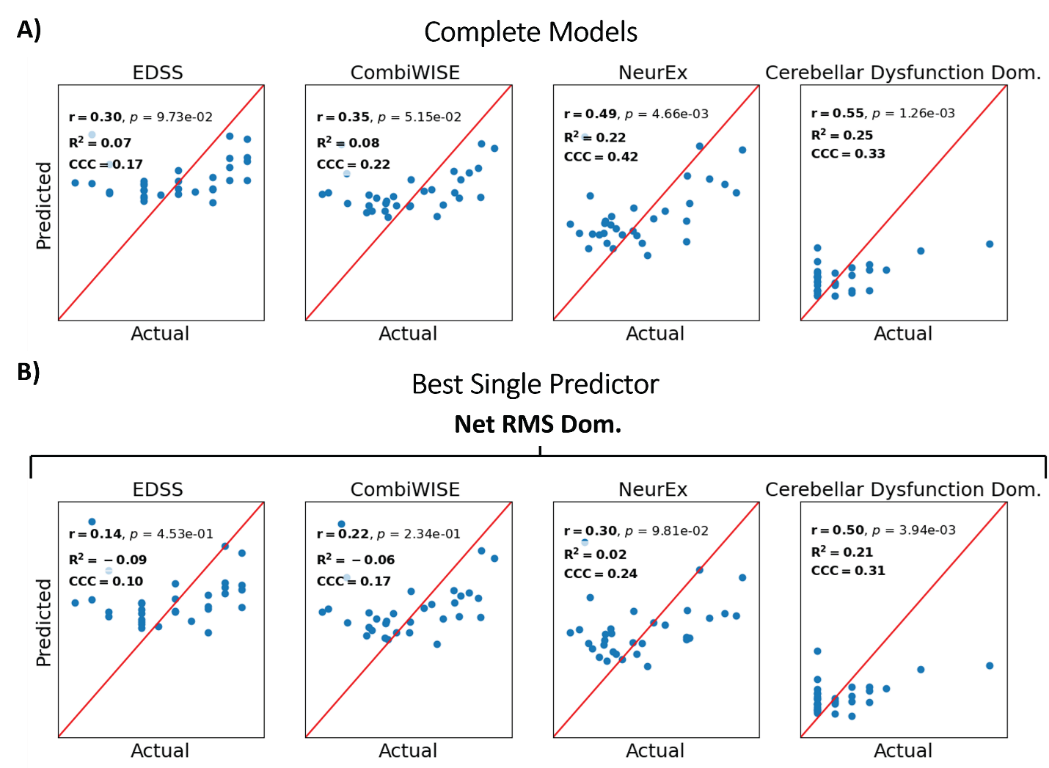

Supplementary Figure 11 | Independent validation results for predictive models cohort compared to the best single predictor for each model outcome (EDSS, CombiWISE, NeurEx^TM^, NeurEx^TM^ Cerebellar Dysfunction in the dominant hand [Dom.]). using Pronator Drift digital biomarkers. Pearson’s r, the coefficient of determination (R^2^), and the concordance correlation coefficient (CCC) were used to evaluate models’ predictive strength for their respective outcomes.

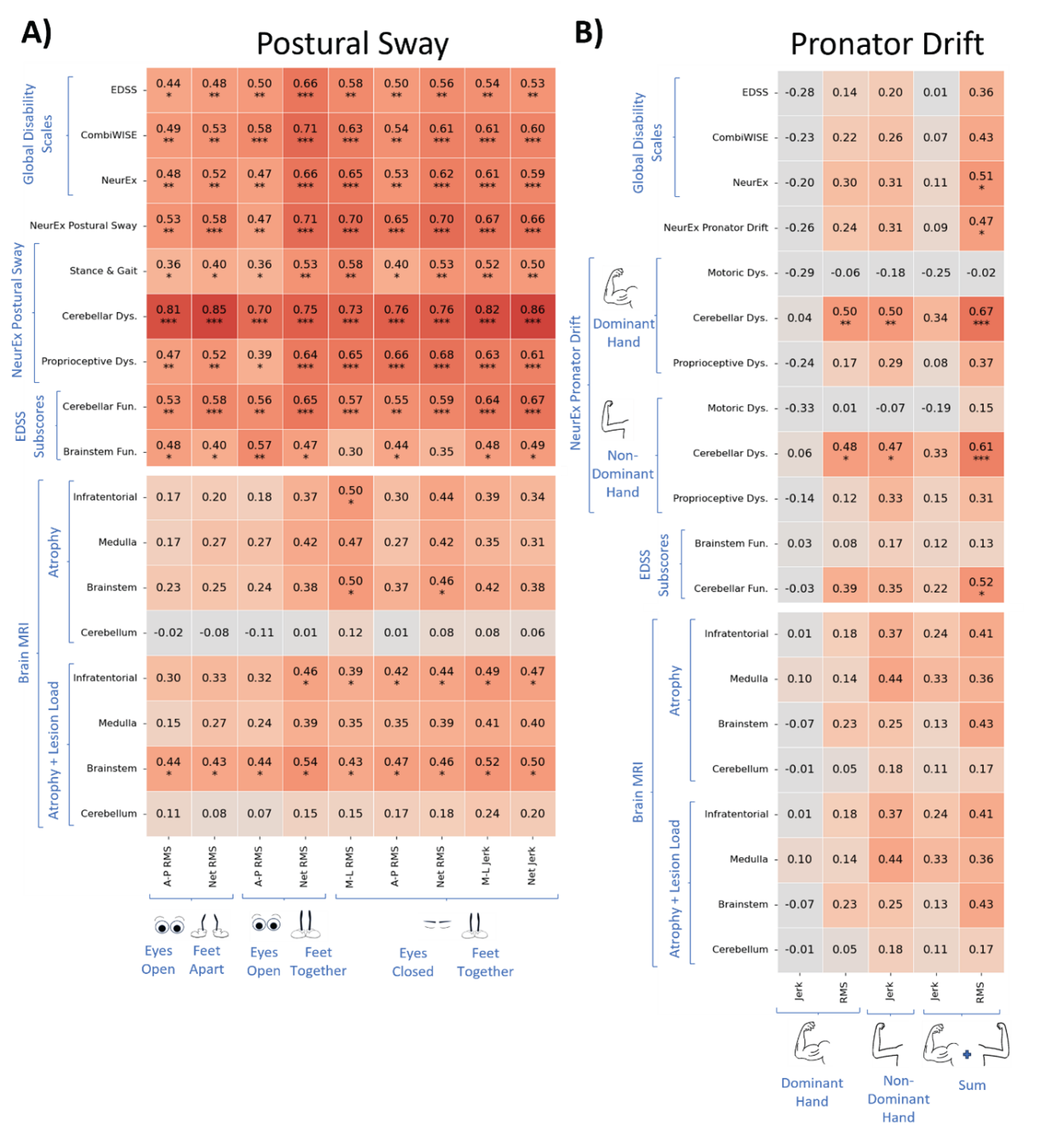

Supplementary Figure 12 | Pearson correlations for the test cohort for between clinical and imaging scores and A) Postural Sway digital biomarkers and B) Pronator Drift digital biomarkers. Correlation p-values were adjusted using the Benjamini-Hochberg False Discovery Rate adjustment with alpha = .05; * indicates a p-value <.05, ** indicates p<.01, *** indicates p<.001. RMS refers to root mean squared acceleration, which captures sway amplitude; Jerk refers to the rate of change of acceleration, which captures sway jerkiness. A-P refers to antero-posterior movement, which captures forward and backward sway; M-L refers to medio-lateral movement, which captures side-to-sway sway; Net combines M-L and A-P acceleration data to capture overall acceleration. “Dys.” is short for “Dysfunction”, “Fun.” is short for “Function”.
